# Untargeted Proteomic Profiling Identifies Candidate Biomarkers for Early Detection of Cardiovascular Disease and Mortality

**DOI:** 10.1101/2025.11.20.25340673

**Authors:** Aleksandra D. Chybowska, Spyros Vernardis, Daniel L. McCartney, Jure Mur, Josephine Robertson, Hannah M. Smith, Archie Campbell, Camilla Drake, Hannah Grant, Poppy Adkin, Matthew White, Christoph B. Messner, Arturas Grauslys, Sergej Andrejev, Charles Brigden, David J. Porteous, Caroline Hayward, Jackie F. Price, Kathryn L. Evans, Aleksej Zelezniak, Markus Ralser, Riccardo E. Marioni

## Abstract

**Background:** The serum proteome can provide valuable insights into the development and progression of diseases. This is particularly important for cardiovascular disease (CVD), a leading cause of death worldwide. In this large-scale cohort study, we employ an untargeted mass-spectrometry-based approach to explore associations between highly expressed proteins, incident CVD (analysed as six individual outcomes and one composite outcome) and all-cause mortality.

**Methods:** The abundances of 439 proteins and protein groups quantified by mass spectrometry in serum were related to incident outcomes in 8,343 Generation Scotland participants (age 40–69 years), who were free of CVD at baseline (n_all_cause_death_=618, n_composite_CVD_=666, follow-up ≤17 years). Cox proportional hazards (PH) models were run before and after adjustment for pre-selected known CVD risk factors. Sex-specific effects were explored. A protein-based risk score for composite CVD outcome was developed using penalised regression.

**Results:** Forty-eight high abundance serum proteins and protein groups were significantly associated with incident CVD and death outcomes (P_Bonferroni_<1.14×10^−4^), including 24 associations not reported in the Open Targets database. Proteins involved in immune and oxidative stress responses were associated with composite CVD (Immunoglobulin heavy variable 3/OR16-9, Hazard Ratio per SD (HR)=0.85 [95%CI 0.79,0.92]) and death (Alpha-1-antitrypsin, HR=1.27 [1.17, 1.38]), while heart failure was linked to proteins playing a role in lipid metabolism (Apolipoprotein A-II, HR=0.70 [0.59, 0.84]) and complement cascade (Complement C1q subcomponent subunit B, HR=1.40 [1.18, 1.66]). Applied to the test set, the proteomic risk score improved 17-year incident CVD prediction over models including age, sex, and nine lifestyle and clinical risk factors (ΔAUC = 0.010, ROC P = 0.013).

**Conclusion:** The highly abundant serum proteome, readily assessed by mass spectrometry, reveals candidate biomarkers for incident CVD and provides predictive value for early risk stratification.

## 2. Introduction

CVD has been among the leading causes of morbidity and mortality worldwide for over 20 years ^1^. Despite progress in diagnosis and treatment, there is an ongoing need to gain further insights into the underlying molecular mechanisms of CVD. This knowledge is essential for improving early diagnosis and developing more effective, targeted therapies.

Currently, there are a number of protein biomarkers known to be associated with acute or chronic CVD, including: 1) cardiac troponin T (cTnT) and I (cTnI) ^2,3^, 2) B-type natriuretic peptide (BNP) and its N-terminal form (NT-pro BNP) ^4^, 3) D-dimer and C-reactive protein (CRP) ^5–7^, and 4) Apolipoprotein A-I ^8^. Elevated levels of cTnT and cTnI are strongly associated with acute coronary syndrome and myocardial infarction ^2,3^. High-sensitivity troponin assays enhance the diagnostic sensitivity for acute coronary syndrome and can also be used for risk stratification. BNP and NT-proBNP are employed to diagnose congestive heart failure ^4^. D-dimer ^7^ and C-reactive protein ^5,6^ are non-specific inflammatory markers that facilitate the diagnosis of thromboembolic conditions and assess cardiovascular risk, particularly in patients with no known cardiovascular disease. Lastly, apolipoprotein A-I is an excellent predictor of HDL metabolism-associated CVD ^8^. While numerous studies have demonstrated the diagnostic and prognostic value of established biomarkers, a key limitation is that many of them reflect downstream or acute manifestations of CVD ^9^. As such, they may be less effective at identifying individuals in preclinical stages of disease or capturing early molecular changes before clinical symptoms emerge. This highlights the need for novel biomarkers capable of detecting early, subclinical pathophysiological processes.

One method of identifying novel biomarkers is by relating circulating protein measures to incident CVD in large cohort studies ^10,11^. Nonetheless, despite this promise, challenges remain. Firstly, CVD is an umbrella term for several inter-related clinical conditions, with atherosclerosis being the common underlying pathology. These conditions include ischaemic heart disease (e.g., angina, myocardial infarction) and cerebrovascular disease (e.g., ischaemic stroke, haemorrhagic stroke, transient ischemic attack). Each sub-condition has its own distinctive risk factor profile and therapeutic strategies, for instance, hypertension may be more critical for stroke ^12^, while cholesterol levels may be useful for coronary heart disease ^13^ though these show considerable overlap. Modelling CVD as a single, composite outcome increases the number of testable cases and can reveal shared biomarkers. However, the heterogeneity of diseases included in the composite outcome can obscure disease-specific associations, and may result in hypothetical biomarkers that are of low specificity.

There are multiple ways of measuring protein concentrations in accessible body fluids. Thus far, most proteomic studies that focused on identifying novel biomarkers of CVD have made use of affinity-reagent based technologies ^14–17^. These targeted technologies can quantify proteins across a wide dynamic range, including many low-abundance components of the circulating proteome. However, because they are limited to a predefined set of proteins, they may miss novel or unanticipated biomarkers. In contrast, mass spectrometry (MS)–based proteomics can be performed in an untargeted fashion, enabling a more comprehensive survey of the proteome. Mass spectrometry is particularly well suited in the quantification of the high abundance serum protein fraction which is enriched in proteins that execute their function in metabolism and immune system, and has thus far proven as the main reservoir of protein biomarkers that eventually made it into clinical use. Experience towards the robust mass spectrometry-based profiling of big cohorts has been gained throughout the recent years_18._

Lastly, men and women differ in their CVD profiles. While young men are typically at a higher risk of developing CVD compared to women of the same age, the risk of CVD in women increases and often surpasses that of men after the menopause, largely due to the loss of the protective effects of oestrogen ^19^. Oestrogen modulates the immune response, balancing pro-inflammatory and anti-inflammatory pathways ^20,21^. By doing so, it helps prevent chronic low-grade inflammation - a key factor in atherosclerosis and other cardiovascular diseases ^22^. Moreover, hormonal contraception use (HCU) has been shown to affect the high abundant proteome ^23^, which may influence the associations with CVD observed in females.

In this work, we use a cost-effective high-throughput MS platform ^24^ to discover candidate biomarkers for the early stages of CVD and gain insights into the molecular aetiology of the studied diseases. The analysis focuses on individuals of European ancestry from the Generation Scotland (GS) cohort. To capture both shared and condition-specific signals, we analysed CVD as a composite outcome - including ischaemic stroke, myocardial infarction, coronary heart disease, and CVD-related death – and as individual events (fatal or non-fatal coronary heart disease, myocardial infarction, heart failure, ischaemic stroke, transient ischaemic attack and CVD death). All-cause mortality was assessed as a separate outcome. Serum proteomic profiling was conducted in 15,818 GS participants, yielding quantitative data on 439 serum proteins and protein groups. Then, we employed Cox regression to link concentration changes in these proteins to CVD and death outcomes. Next, we analysed sex-specific effects of proteins on CVD risk and the influence of established lifestyle and health risk factors on these associations. Finally, we derived a proteomic risk score for the composite CVD outcome and evaluated its utility for 17-year risk prediction.

## Methods

### 2.1. Generation Scotland

Generation Scotland (GS) is a family-based cohort of individuals aged 18 to 98 from 7,000 family groups spread throughout Scotland ^25^. Recruitment of study participants occurred between 2006 and 2011. Eligible individuals were invited to attend a clinic, where their clinical and physical characteristics were measured following a standardized protocol. In total, 24,088 participants completed a health questionnaire. Fasting blood samples were collected from 21,521 individuals using a standard operating procedure. All participants provided written informed consent for research. The study received ethical approval from the National Health Service Tayside Committee on Medical Research Ethics (REC reference number: 05/S1401/89). The GS dataset is not publicly available as it contains information that could compromise participant consent and confidentiality. However, the data, research materials, and analytical methods will be made accessible to other researchers for the purpose of replicating the findings. Access will be granted upon successful project application to the GS Access Committee and obtaining ethical approval for accessing linked health data from NHS Scotland. Instructions for accessing GS data can be found at https://www.ed.ac.uk/generation-scotland/for-researchers/access; the GS Access Request Form can be downloaded from this site. All code used in the analyses is available with open access at the following Github repository: https://github.com/aleksandra-chybowska/MS_proteins_and_CVD.

### 2.2. Materials

For the mass spectrometric proteome analysis study, the following chemicals and items were used: acetonitrile (ACN, Thermo Fisher, 10001334), ammonium bicarbonate (ABC, Thermo Fisher, 15645440), C18 96-well plates (BioPureSPN Macro 96-well, 100 mg PROTO 300 C18, HNS S18V-L), dithiothreitol (DTT, Sigma-Aldrich, 43815), formic acid (FA, Thermo Fisher Scientific, 85178), iodoacetamide (IAA, Sigma-Aldrich, I1149), methanol (MeOH, Thermo Fisher, 10767665), trypsin (Promega, Sequence Grade, V5117), urea (Sigma-Aldrich, 1084870500) and water (Thermo Fisher, 10505904).

### 2.3. Sample Preparation for Serum Proteomics

The sample preparation process has been described previously ^24,26^. Briefly, 5 μL of serum samples were added to a solution of 50 μL of 8 M urea and 0.1 M ammonium bicarbonate at pH 8.0 to denature the proteins. Subsequently, the proteins were reduced using 5 μL of 50 mM dithiothreitol for 1 hour at 30 °C and alkylated with 5 μL of 100 mM iodoacetamide for 30 minutes in the dark. The sample was then diluted with 340 μL of 0.1 M ammonium bicarbonate to a concentration of 1.5 M urea. The next step was the trypsinisation of the proteins and 200 μL of the solution was used, and the proteins were digested overnight with trypsin (12.5 μL, 0.1 μg/μL) at 37 °C at a 1/40 trypsin/total protein ratio. The digestion was quenched with the addition of 25 μL of 0.1% v/v FA. The peptides were cleaned up with C18 96-well plates, eluted with 50% v/v ACN, dried by a vacuum concentrator (Eppendorf Concentrator Plus), and redissolved in 50 μL of 10% v/v FA for processing by liquid chromatography (LC)-MS.

### 2.4. Liquid Chromatography and Mass Spectrometry Acquisition

An Agilent 1290 Infinity II system (Agilent Technologies) was used for liquid chromatography, and it was connected to a TripleTOF 6600 mass spectrometer (SCIEX). 2 μg of total peptides were injected and separated using a Luna Omega LC Column (1.6u PS C18 100A, 30 x 2.1 mm (Phenomenex)) over a 3-minute gradient. A linear gradient was utilised, starting from 1% B and reaching 40% B over 3 minutes (Buffer A: 0.1% v/v FA; Buffer B: ACN/0.1% v/v FA) with an 800 μL/min flow rate. When the gradient reached 40%, the washing and re-equilibration steps were carried out in the following manner: B was increased from 40% to 80% over 0.5 min, followed by 80% B for 0.2 min, and then decreased from 80% to 3% B over 0.1 min. Re-equilibration at 3% B was maintained for 1 min until the next injection.

For the sample acquisition, we used a Scanning SWATH method ^27^. The mass spectrometer was operated at high-sensitivity mode. The source conditions were as follows: source gas one at 15 psi, source gas two at 20 psi, curtain gas at 25 psi, temperature at 0 °C, IonSpray floating voltage at 5,500 V, and declustering potential at 80 V. Rolling collision energies were determined using the following equation: CE=0.034×𝑚/𝑧+2, where m/z represents the centre of the scanning quadrupole bin. The Scanning SWATH parameters were as follows: total cycle time 0.69 s, transmission window width 10 Da, Sample duration 3.103 min, precursor range 450-850 Da, fragment range 100-1500 Da and effective accumulation time 16.90 ms.

### 2.5. Mass Spectrometry Data Processing, Batch Correction and Quality Control

Raw data was analysed by DIA-NN ^28^ as described previously ^24^. DIA-NN (version 1.8.12) was run in Robust LC (high precision) quantification mode, using the default parameter set. Identification was performed using a previously generated spectral library ^29^, which was refined using DIA-NN, as described before ^24^. During all steps, precursor false discovery rate (FDR) filtering was set to 1%. **Supplemental Figures 1** and **2** show the distribution of identified precursors.

Postprocessing was carried out in R v4.3.1 ^30^. The data was filtered at 1% gene group q-value and only samples with minimum precursor identifications of 2000 were included in the analysis. The precursors were filtered to have at least 80% prevalence in QC samples. Within-batch signal drift correction was performed by fitting a linear model to the repeat injections of pooled QC samples and using the model to correct the remaining samples (adapted from ^31^). The between-batch correction was done using the “limma” v3.54.2 ^32^ linear batch-correction algorithm. Prior to running subsequent analyses, all proteins were annotated (**Supplemental Table 1, Supplemental Methods**) and their abundances were rank-based inverse normalised (n=15,818).

### 2.6. Selection of Covariates for Survival Analysis

Covariates of our Cox PH models included known CVD risk factors, such as age, sex, average systolic blood pressure, total cholesterol, HDL cholesterol, smoking status, presence of rheumatoid arthritis, diabetes status, years of education, and socioeconomic deprivation (measured by the Scottish Index of Multiple Deprivation, SIMD). The assessment of these variables is explained in detail in the **Supplemental Methods** section. In line with the approach implemented by the authors of SCORE2 clinical risk predictor ^33^, all individuals aged less than 40 years and more than 69 years were excluded from the analysis. Average systolic blood pressure, log transformed pack years of smoking, HDL cholesterol and total cholesterol levels were trimmed of outliers (points beyond 4 standard deviations (SDs) of the mean). Body mass index (BMI) was log transformed and filtered to values between 18 and 50 kg/m^2^. Participants with missing covariate data were excluded from the dataset, leaving a sample size of n=8,343 (**Supplemental Figure 3**).

Given that recent studies suggest a link between the circulating proteome and HCU in females^23,34^, we examined this relationship using linear regression (**Supplemental Methods**). As significant associations between the studied proteins and HCU were detected (**Supplemental Table 2**), we decided to further adjust our models for contraception use.

To assess multicollinearity among the covariates, we generated a Spearman correlation matrix (**Supplemental Figure 4**) and calculated the variance inflation factor (VIF) (**Supplemental Table 3**). As all variable pairs had Spearman’s r < 0.6 and the VIF values for all variables were less than 2, we did not exclude any of the covariates from the analysis due to multicollinearity.

### 2.7. Outcome Definitions for Survival Analysis

CVD (hospitalised cases only) and all-cause mortality data were identified through linkage to NHS data and death records. Incident cases were ascertained over a follow up period of up to 17 years. The outcomes of interest were divided into three categories: a) individual CVD events, which included fatal or non-fatal coronary heart disease, myocardial infarction, heart failure, ischaemic stroke, transient ischaemic attack, and CVD death, b) a composite CVD outcome, which included diseases considered in a previous GS publication by Welsh *et al*. ^35^ (coronary heart disease, ischaemic stroke, myocardial infarction and CVD death), and c) all-cause death. All disease outcomes were defined as per CALIBER/HDR UK ^36^ consensus definitions. **Supplemental Table 4** details International Classification of Diseases, 10th Revision (ICD10) codes included in each sub-category of outcomes. Only the first occurrences of events were considered per sub-category. More information about intersections of events within GS can be found in **Supplemental Figure 5**. CVD death was defined in line with Welsh *et al*. ^35^ (**Supplemental Table 4**).

### 2.8. Survival Analysis

The study employed Cox PH regression implemented in Python ^37^ version 3.10.13 using lifelines library ^38^ version 0.27.8 to investigate the relationship between individual protein abundances and the studied outcomes. CVD cases included individuals diagnosed after baseline who subsequently died, as well as those who remained alive after diagnosis. Prevalent cases were excluded from the analysis. Controls were censored at the end of the follow-up period (August 2023, up to 17 years of follow up) or at the time of death.

For each protein, two types of models were considered: a) basic models adjusted for age and sex, and b) fully-adjusted models including age, sex, average systolic blood pressure, total cholesterol, HDL cholesterol, smoking (pack years), rheumatoid arthritis, diabetes, years of education, SIMD score, and contraceptive use. Protein abundance was included in all models as the primary independent variable.

Sex-specific effects of proteins on the outcome risk were studied by incorporating a sex by protein abundance interaction term (protein*sex) to fully adjusted models (b) and plotted using “plot_partial_effects_on_outcome” function from lifelines library ^38^. This function uses previously fitted Cox model to visualise the effect of varying one or more covariates on the predicted survival probability.

Proteins significantly associated with the studied outcomes in fully-adjusted models (b) were further analysed in R version 4.3.1 ^30^. The linearity assumption was assessed by examining deviance residuals using ggcoxdiagnostics function from the survminer package ^39^ version 0.4.9. Full models were re-run using coxme library ^40^ version 2.2-20 with a kinship matrix fitted as a random effect to adjust for relatedness.

All models were adjusted for multiple testing using a Bonferroni correction applied in a disease-specific manner (P_Bonferroni_=0.05/439). Given its conservative nature, we compared the findings after applying an FDR-correction (P_FDR_<0.05).

### 2.9. Statistical Attenuation After Covariate Adjustment

An additional series of Cox PH regressions were run to model time to a composite CVD outcome for proteins that were significant in the basic model but not in the fully adjusted model. The analysis aimed to identify proteins that initially associated with CVD outcomes but failed to meet the threshold for statistical significance after adjusting for additional variables. The basic model was adjusted for age, sex, and individual protein abundance. Subsequently, additional covariates - average blood pressure, total cholesterol, HDL cholesterol, smoking, rheumatoid arthritis, diabetes, years of education, SIMD score, and contraceptive use - were independently incorporated into the basic model. The impact of the covariates was assessed by comparing the protein’s significance to that from the basic model.

Using a similar approach, associations from the basic model were compared with those from a model additionally adjusted for baseline use of cardiovascular drugs, to assess potential confounding by treatment. Details of medication use assessment are provided in the **Supplemental Methods.**

### 2.10. Proteomic CVD Risk Score

A proteomic risk score for a composite CVD outcome was developed using elastic net regression. The proteomic dataset with incident CVD data collected over 17-year follow-up period (n=8343) was split into training and test sets. Individuals unrelated to each other and to all individuals in the training set (“singletons”) were selected based on family ID variable and assigned to the test set (n=2660 individuals, 229 events). The dataset containing remaining participants was used for training (n=5683 individuals, 437 events). Protein abundances were rank-based inverse normal transformed and scaled (mean=0, SD=1) separately within training and test datasets.

In the training set, deviance residuals from an age- and sex-adjusted Cox model of composite CVD were used as the outcome. Linear elastic net regression (glmnet v4.1) ^41^ was fitted with 10-fold cross-validation stratified by family ID to prevent related individuals from splitting across folds. Proteins with non-zero coefficients were retained, and their coefficients were used to compute a weighted linear combination of protein abundances (the proteomic score). In the test set, scores were calculated using the selected coefficients. Four nested Cox PH models were evaluated for each outcome: (a) minimally adjusted (age, sex), (b) model (a) + proteomic score, (c) extended model including age, sex, and nine lifestyle and clinical risk factors (average systolic blood pressure, total cholesterol, HDL cholesterol, smoking pack years, rheumatoid arthritis status, diabetes status, educational attainment, SIMD, contraception use), and (d) model (c) + proteomic score. Model discrimination was assessed over 17 years of follow-up using C-index and area under the Receiver operating curve (AUC). Improvement from adding the proteomic score was quantified by comparing Receiver operating (ROC) curves using pROC ^42^ R package.

## 3. Results

### 3.1. Baseline Characteristics

Detectable abundances of 133 (30.3%) proteins and 306 (69.7%) ‘protein groups’ were found in 15,818 GS individuals. Protein groups are defined as mass spectrometry results that can be assigned to multiple proteins due to shared amino-acid sequences. Among identified protein groups, 199 (65.0%) originated from the same gene. Here, we refer to them as gene-derived protein groups (e.g., haptoglobin (G1) and haptoglobin (G2)). Of these, 61 (30.7%) protein groups occurred only once in the protein group subset of the dataset, such as selenoprotein P (G). Remaining protein groups (n=107, 35.0%) are labelled with a symbol “(PG)”.

In our analysis sample of 8,343 participants (**Table 1**, detailed characteristics by outcome are provided in **Supplemental Table** 5), individuals who developed composite CVD (n=666) were more likely to be male and generally exhibited a more adverse risk factor profile. This included older age, elevated body mass index, lower HDL cholesterol and higher average systolic blood pressure. They also smoked more, spent less time in full-time education, were more likely to take cardiovascular medications, and had a higher prevalence of baseline diabetes or rheumatoid arthritis.

**Table 1.** Baseline Demographic Characteristics of the Study Cohort. *P* values correspond to independent sample *t* tests for continuous variables with normal distribution (reported as average ± SD), Mann–Whitney *U* test for continuous variables with skewed distribution (reported as median [quartile 1, quartile 3], or χ^2^ test for categorical variables (reported as n [%]). BMI indicates body mass index; CVD, cardiovascular disease; HDL, high-density lipoprotein; SBP, systolic blood pressure; and SIMD, Scottish Index of Multiple Deprivation. BMI and smoking were log-transformed prior to running the statistical comparisons.

| *Characteristic* | *No Composite CVD*<br>*(n = 7677)* | *Composite CVD*<br>*(n = 666)* | *P Value* |
| --- | --- | --- | --- |
| *Age, years* | 53.7 (7.5) | 58.1 (6.7) | <0.001 |
| *Male sex, n (%)* | 3041 (39.6) | 428 (64.3) | <0.001 |
| *BMI, kg/m<sup>2</sup>* | 27.15 (4.83) | 28.60 (5.31) | <0.001 |
| *SIMD, rank* | 4593 [2708, 5554] | 4178 [2213, 5483] | 0.002 |
| *Diabetes, n (%)* | 222 (2.9) | 68 (10.2) | <0.001 |
| *Rheumatoid arthritis, n (%)* | 142 (1.8) | 18 (2.7) | 0.164 |
| *Medication use, n (%)* | 1483 (19.3) | 278 (41.7) | <0.001 |
| *Smoking, pack years* | 0 [0, 13] | 5 [0, 34] | <0.001 |
| *SBP, mm Hg* | 133.8 (17.3) | 140.6 (18.4) | <0.001 |
| *Total cholesterol, mmol/L* | 5.4 (1.0) | 5.3 (1.2) | 0.171 |
| *Non-HDL cholesterol, mmol/L \** | 3.9 (1.0) | 4.0 (1.1) | 0.020 |
| *HDL cholesterol, mmol/L* | 1.4 [1.2, 1.7] | 1.3 [1.1, 1.6] | <0.001 |
| *Full-time education\** | 4 [3, 6] | 4 [3, 5] | <0.001 |

\* The medications variable indicates treatment with cardiovascular medications. Non-HDL cholesterol was calculated as total cholesterol minus HDL cholesterol concentration. Full-time education is a categorical variable, reported here as the median [quartile 1, quartile 3] for data privacy reasons. The categories are defined as follows: 0 (0 years), 1 (1–4 years), 2 (5–9 years), 3 (10–11 years), 4 (12–13 years), 5 (14–15 years), 6 (16–17 years), 7 (18–19 years), 8 (20–21 years), 9 (22–23 years), and 10 (24 or more years).

### 3.2. Protein Abundances are Associated with CVD Outcomes and Death

We tested whether abundances of 439 proteins and protein groups were associated with risk of cardiovascular outcomes and death over 17 years of follow-up. Risk was modelled using Cox PH regression, with one protein considered per model (**Table 2, Supplemental Tables 6** and **7**).

**Table 2.** The Number of Proteins Associated with CVD and Death. . Mean time to event is reported together with its SD. The basic models were adjusted for age, sex, and the abundance of the studied protein. The fully-adjusted models included these factors as well as additional covariates: average blood pressure, total cholesterol, HDL cholesterol, smoking status, rheumatoid arthritis, diabetes, years of education, SIMD score, and contraceptive use. Significant P values were corrected for multiple testing via the Benjamini-Hochberg method (P_FDR_<0.05) and separately using the Bonferroni method (P_Bonferroni_<1.14×10^−4^).

| Event | Number of events | Time to Event (years) | Significant proteins (n) |  |  |  |
| --- | --- | --- | --- | --- | --- | --- |
| | | | Basic ($P < 0.05$) | Full ($P < 0.05$) | Full ($P_{\text{FDR}}$) | Full ($P_{\text{Bonferroni}}$) |
| **Composite CVD** | 666 | 7.6 (4.1) | 194 | 109 | 42 | 9 |
| **CVD Death** | 245 | 9.6 (4.1) | 164 | 109 | 46 | 4 |
| **All Cause Death** | 618 | 9.2 (4.1) | 179 | 122 | 70 | 28 |
| **Coronary Heart Disease** | 329 | 7.2 (3.8) | 164 | 56 | 0 | 0 |
| **Myocardial Infarction** | 233 | 7.1 (3.9) | 140 | 48 | 0 | 0 |
| **Heart Failure** | 140 | 7.8 (3.9) | 126 | 81 | 27 | 7 |
| **Ischaemic Stroke** | 99 | 8.3 (3.6) | 53 | 27 | 0 | 0 |
| **Transient Ischaemic Attack** | 85 | 7.6 (3.7) | 67 | 34 | 0 | 0 |

In the basic models, which were adjusted for age, sex, and the abundance of the specific protein, 243 proteins and protein groups showed significant associations with the health outcomes after Bonferroni correction for multiple testing (P_Bonferroni_<1.1×10^−4^). Out of these, 29 did not satisfy the PH assumption (P_local_test_<0.05) and were excluded from further analysis. 116 proteins were linked to multiple outcomes.

In the full models, which were further adjusted for pre-selected cardiovascular risk factors and the use of hormonal contraception (as detailed in the **Methods** section), the number of significant associations with health outcomes decreased to 48 (P_Bonferroni_<1.1×10^−4^). Among these, 41 met the PH assumption, with seven proteins linked to multiple outcomes. Twenty-four of the 41 associations between proteins and events had not been previously reported in the Open Targets database ^43^, which integrates a wide range of data sources on target-disease relationships. Adjusting for kinship did not affect the findings. Further details on previously unreported associations and the impact of adjusting for relatedness are provided in **Supplemental Table 8**.

Figure 1 illustrates Bonferroni-significant associations after adjusting for risk factors in the full models. Multiple patterns were observed for these findings.

**Figure 1.**
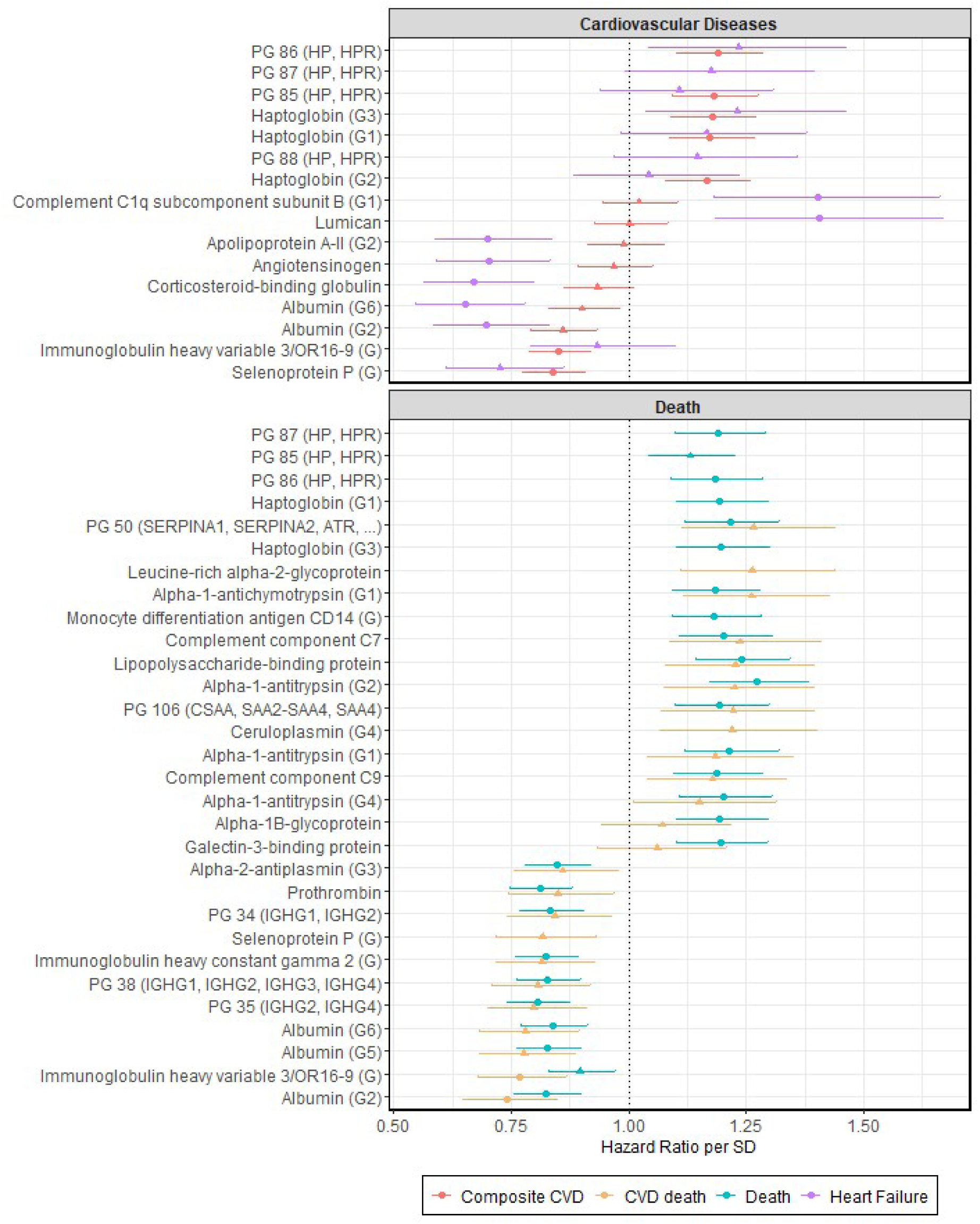
Association Between Mass Spectrometry Protein Abundances, Death, and Cardiovascular Outcomes. The results are based on fully-adjusted models. Only the associations that met the Cox Proportional Hazards assumption are shown. Proteins and protein groups significant after applying Bonferroni correction are marked with circles, while non-significant associations are marked with triangles. Error bars represent 95% confidence intervals.

**Figure 2.**
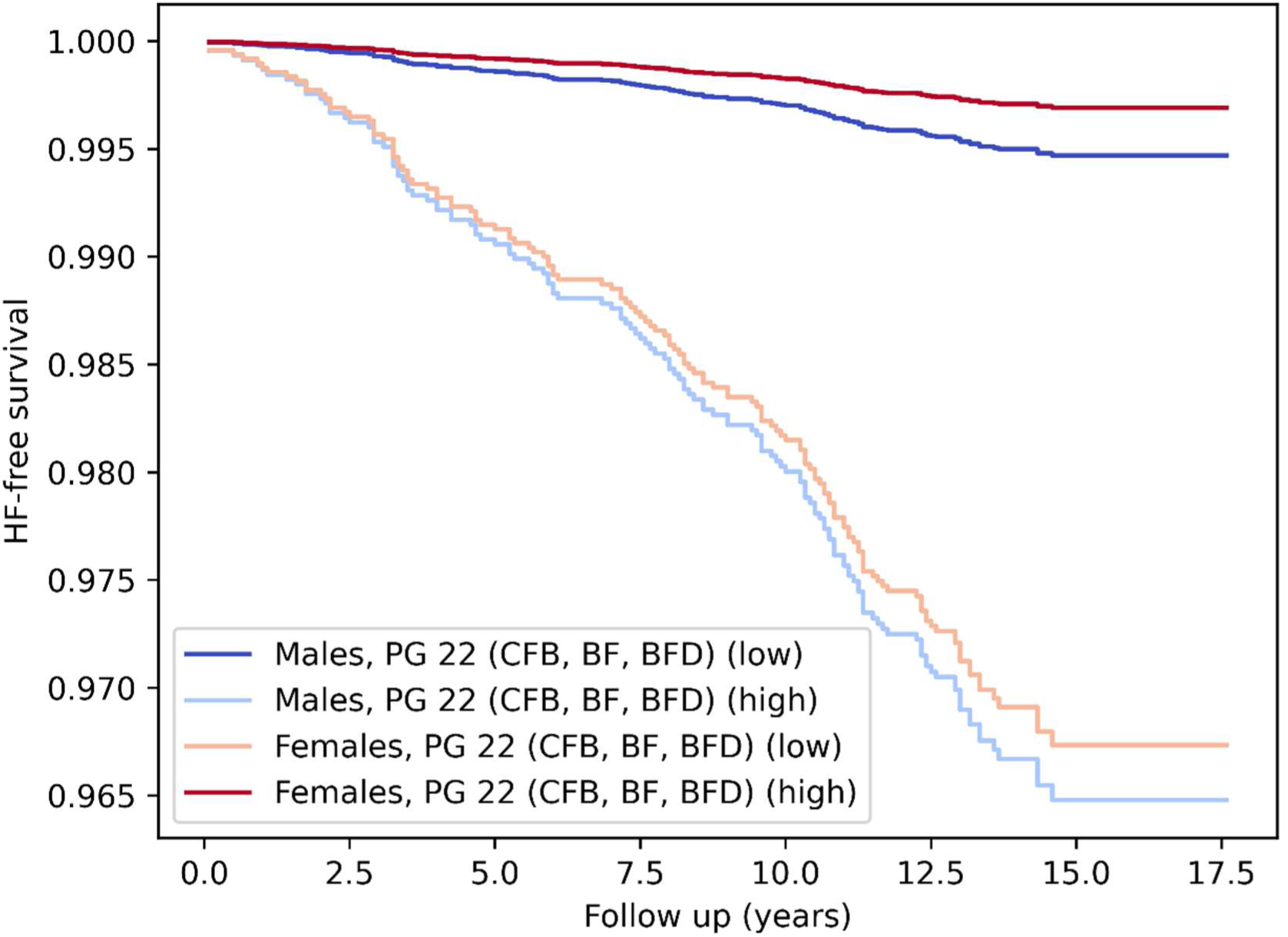
Predicted Sex-specific Effects of PG 22 (CFB, BF, BFD) on Heart Failure Risk. This protein group (containing complement factors B and C2) was associated with a decreased risk of heart failure in females, but with an increased risk in males. The high and low categories represent 3 SDs above and below the mean abundance of PG 22, respectively.

Firstly, increased abundances of haptoglobin isoforms (protein groups G1 and G3, HR range 1.17-1.18, P<1.1×10^−4^) were associated with an increased hazard (HR>1) of composite CVD. In contrast, isoforms of selenoprotein P (G) were associated with a reduced hazard (HR<1) of CVD (HR=0.84 [0.78; 0.91], P=9.1×10^−6^) and all-cause mortality (HR=0.83 [0.77, 0.90], P=9.7×10^−6^). The association between Immunoglobulin heavy variable 3/OR16-9 (G) and both CVD (HR=0.85 [0.79, 0.92], P=4.1×10⁻⁵) and CVD-related death (HR=0.77 [0.68, 0.87], P=2.3×10⁻⁵) was not reported in the Open Targets database.

Secondly, Lumican (HR=1.41 [1.18,1.67], P=1.1×10^−4^) was associated with an increased hazard of heart failure, while Angiotensinogen (HR=0.70 [0.59, 0.83], P=4.3×10^−5^) was linked to a reduced hazard. The association between Angiotensinogen and heart failure was not driven by confounding due to HCU (P_HCU_ > 0.05). Previously unreported protein associations with heart failure risk included Apolipoprotein A-II (G2) (HR=0.70 [0.59, 0.84], P=7.5×10^−5^), Corticosteroid-binding globulin (HR=0.67 [0.57, 0.80], P=5.2×10^−6^) and Complement C1q subcomponent subunit B (G1) (HR=1.40 [1.18, 1.66], P=1.1×10^−4^).

The three most statistically significant associations with an increased risk of all-cause mortality were unreported in Open Targets and included Alpha-1-antitrypsin (G2) (HR=1.27 [1.17, 1.38], P=1.2×10^−8^), Lipopolysaccharide-binding protein (HR=1.24 [1.14, 1.35], P=2.8×10^−7^) and Leucine-rich alpha-2-glycoprotein (HR=1.24 [1.14, 1.34], P=4.7×10^−7^). Among the three proteins most significantly associated with a reduced risk of all-cause mortality, two were unreported - PG 35 (IGHG2, IGHG4) (HR=0.81 [0.74, 0.87], P=2.2×10⁻⁷) and Immunoglobulin heavy constant gamma 2 (G) (HR=0.82 [0.76, 0.89], P=2.3×10⁻⁶) - while one, Prothrombin (HR=0.81 [0.75, 0.88], P=6.3×10⁻⁷), was a previously known association.

Finally, while associations with coronary heart disease, myocardial infarction, ischaemic stroke, and transient ischaemic attack did not remain significant after correcting for multiple testing, proteins linked with: 1) coronary heart disease and myocardial infarction, 2) ischemic stroke and transient ischemic attack, as well as 3) CVD death and all-cause death exhibited similar effect directions and magnitudes across basic and fully-adjusted models (**Supplemental** Figures 6, 7 and 8).

### 3.3. Sex-specific Effects

We also investigated sex-specific effects of proteins on the risk of the studied health outcomes. As before, we ran 439 Cox PH regressions, but now incorporating an interaction term (protein*sex). This analysis revealed a protein group with significant sex-specific effects on the risk of heart failure (P_Bonferroni_<1.1×10^−4^). A protein group containing complement factors B and C2 (HR_protein*sex_=2.04 [1.45, 2.87], P=4.1×10^−5^) was associated with a decreased risk of heart failure in females, but increased the risk in males (Figure 2).

### 3.4. Risk Factors Driving Associations with Incident CVD Outcome

55 protein-incident CVD associations were statistically significant at a Bonferroni threshold (P<1.1×10^−4^) in the basic model but not the fully-adjusted model. To determine which variables were potentially attenuating the associations (in terms of statistical significance), the basic model was augmented with each covariate in isolation (more details, including applied formulas, are available in **Supplemental Table 9**). The following number of associations with proteins were attenuated after the addition of: HDL cholesterol (24), diabetes status (13), pack years (11), years of education (9), SIMD score (8), average systolic blood pressure (5), rheumatoid arthritis (1). Of the 55 proteins, 17 have not previously been reported in the Open Targets database ^43^ in relation to CVD. Previously unreported biomarkers (n=9) included proteins such as Alpha-2-antiplasmin (G1), Immunoglobulin heavy variable 3-7 and PG 107 (VTN) (**Supplemental Table 9**).

Two proteins (Apolipoprotein B-100 and Apolipoprotein B-100 (G)) were not significant in the basic model of composite CVD but become significant at P_FDR_<0.05 after adjusting the basic model for cardiovascular medication use. A full list of associations reaching P<0.05 in the age-, sex-, and medication-adjusted model is provided in **Supplemental Table 10**.

### 3.5. Proteomic CVD Risk Score

To assess whether mass spectrometry–derived protein abundances add predictive value beyond established CVD risk factors, we developed a proteomic CVD risk score. An elastic net regression model was trained on 5683 individuals with 437 events, selecting 50 of 439 measured proteins (coefficients in **Supplemental Table 11**).

The score was tested in a hold-out set of 2660 individuals (n=229 events) using four 17-year Cox proportional hazards models of increasing complexity. The hold-out set were both unrelated to each other and to all individuals in the training set, based on the family ID variable in GS. The minimally adjusted model (age and sex) achieved an AUC of 0.689 (C-index=0.685). Adding the proteomic score improved performance to AUC=0.729 (C-index=0.725), with the score showing independent predictive value (HR=1.47 [1.31–1.65], P=4.1×10^−11^). The extended model with age, sex, and the nine lifestyle and clinical risk factors reached an AUC of 0.741 (C-index=0.737), while further inclusion of the proteomic score yielded the best discrimination (AUC=0.751, C-index=0.745). The proteomic score remained a significant predictor beyond established risk factors (HR=1.25 [1.09–1.44], P=0.002). Differences in ROC curves were significant for both the minimally adjusted model with vs. without the proteomic score ( Δ AUC=0.040, Δ ROC P=4.4×10^−5^) and the extended model with vs. without the score ( Δ AUC = 0.010, ROC P = 0.013).

## 4. Discussion

We conducted a large-scale, unbiased study using untargeted mass spectrometry to analyse the circulating proteome and its association with CVD and mortality. This approach focused on high-abundance proteins, which are critical to metabolism and immune function and have historically yielded successful biomarkers. We identified 48 proteins and protein groups that improve CVD and mortality risk prediction beyond traditional factors, with associations detectable up to 17 years before the event. Of these, 24 protein-disease links were not present in Open Targets database. Additionally, we explored sex-specific effects of the complement system in heart failure and developed a proteomic risk score that improved the prediction of incident CVD beyond established risk factors. These findings could inform the development of novel biomarkers for early CVD detection and personalized prevention strategies.

Our results align with the immuno-inflammatory theory of CVD, which suggests that inflammation and immune system dysregulation play a central role in the development and progression of CVD. Proteins associated with an increased hazard of the composite CVD outcome included haptoglobin, an acute-phase reactant whose concentrations fluctuate significantly during inflammation. ^44^. Haptoglobin is also responsible for the clearance of toxic free haemoglobin released during red blood cell breakdown. By binding free haemoglobin, it mitigates tissues damage and oxidative stress, contributing to cardiovascular health ^44^. Similarly, a selenium transporter called Selenoprotein P works to support the immune system and reduce oxidative stress ^45^. It associated with a decreased risk of CVD. Additionally, isoforms of Immunoglobulin heavy variable 3/OR16-9 were associated with a decreased risk of CVD and were unreported in the Open Targets database.

Inflammation-related proteins were also identified in associations with all-cause and CVD death. While alpha-1-antitrypsin ^46^ modulates the immune response and protects tissues from inflammatory damage, Lipopolysaccharide-binding protein ^47^, and Leucine-rich alpha-2-glycoprotein ^48^ are acute phase reactants expressed in ongoing inflammation. In contrast, isoforms of immunoglobulin heavy constant gamma 2 are subclasses of IgG antibodies, which are critical components of the adaptive immune system. The inverse association between their abundance and all-cause mortality has not been reported before.

Proteins associated with heart failure seemed to track the progression of CVD. Apolipoprotein A-II is the second most abundant apolipoproteins of HDL. Although its role in HDL function and metabolism has been debated ^49^, epidemiological studies showed that it is inversely associated with risk of future coronary artery disease ^50^, possibly through its involvement in reverse cholesterol transport and removal from arteries ^51^. Component C1q has been implicated in the initiation and progression of atherosclerotic plaques and promoting plaque instability ^52^, whereas Lumican ^53^ is involved cardiac remodelling ^54^.

The negative association of angiotensinogen with heart failure was unexpected. It did not reflect confounding by hormonal contraception ^23^. Angiotensinogen is a part of the renin-angiotensin-aldosterone system, which regulates blood pressure ^55^. Briefly, when the pressure drops, renin is released. Renin converts angiotensinogen to angiotensinogen I, which is then converted to angiotensin II. Angiotensin II stimulates aldosterone secretion in the adrenal glands, leading to salt and water reabsorption by the kidneys and constriction of small arteries, thereby increasing blood pressure. The protective effect of angiotensinogen may represent an adaptive response to the heart’s diminished capacity to meet the body’s blood flow demands during heart failure ^56^. This adaptation involves the upregulation of angiotensin-converting enzyme 2 (ACE2), which converts angiotensin II into angiotensin 1–7 ^56^. Angiotensin 1–7 exerts vasodilatory effects, counterbalancing the actions of angiotensin II ^57^. The upregulation of the ACE2/angiotensin 1–7 axis in heart failure serves as a compensatory response to mitigate the detrimental effects of an overactive renin-angiotensin system, thereby preserving cardiac function ^57^.

Our results suggest that the differential risk of heart failure between males and females is accounted for by protein groups associated with complement system. Specifically, complement factors B and C2 were associated with a decreased risk in females but an increased risk in males. Further research is warranted to explore the underlying mechanisms driving these sex-specific associations.

Whilst conducting Mendelian randomisation analyses was beyond the scope of this study, we have addressed this question as part of a broader genome-wide association study of the mass spectrometry–derived proteins ^58^. That work identified three proteins - Apolipoprotein B, Apolipoprotein E, and Apolipoprotein - as causally linked to coronary artery disease. In our current analyses, Apolipoprotein and Apolipoprotein E showed significant associations at P_FDR_<0.05 in the basic models of composite CVD and CVD death, respectively, but these associations attenuated after accounting for established risk factors, highlighting the influence of conventional clinical variables. Interestingly, Apolipoprotein B was not significant in the basic model of composite CVD, yet became significant at P_FDR_<0.05 after adjusting for cardiovascular medication use, suggesting that therapeutic interventions may mask its true contribution to CVD risk. These observations underscore the importance of considering treatment effects when interpreting proteomic associations.

Our findings demonstrate that the newly developed proteomic risk score provides independent predictive value for CVD up to 17 years before the event. These results are consistent with previous studies using targeted affinity-based panels such as Olink and SOMAScan ^14–17^, which have also highlighted the utility of circulating proteins in risk prediction. By applying an untargeted mass spectrometry approach, our study captured proteins not typically included in targeted panels, which may represent novel contributors to long-term risk prediction. Furthermore, while most prior studies have concentrated on 10-year prediction horizons, our results suggest that proteomic profiling holds promise for identifying at-risk individuals much earlier in the disease course.

Study strengths include utilising one of the world’s largest mass spectrometry proteomics cohort studies, which also contained hundreds of incident CVD and death events. Many of the associations observed have not, to our knowledge, been previously studied in the context of CVD. Our unbiased approach, including rarely studied protein isoforms, adds depth and breadth to the underlying biology of CVD risk. The long follow-up period further supports early detection and risk assessment.

Despite these strengths, several limitations must be acknowledged. Firstly, the study cohort consists exclusively of individuals of European ancestries residing in Scotland, which may limit the generalisability of our findings to other populations. Secondly, while highly scalable and affordable, our mass spectrometry approach tends to measure the more abundant fraction of the proteome, excluding some known biomarkers such as cardiac troponin or C-reactive protein. Thirdly, in cases where peptides could not be uniquely assigned to a single protein, PGs were created. Each PG may contain several proteins that share peptides, meaning it is not always possible to identify which protein drives the measured signal. In addition, we identified previously unreported associations by cross-referencing with the Open Targets database. However, Open Targets is not fully comprehensive and may not include all published or emerging evidence. Finally, while external replication would strengthen the cardiovascular relevance and robustness of our findings, no comparable mass spectrometry-based dataset currently exists to support direct validation.

In conclusion, our findings demonstrate that circulating proteins and their isoforms are associated with incident CVD and mortality up to 17 years before the event, independent of traditional risk factors. By employing a mass spectrometry-based approach, we identified previously unreported associations that may have been overlooked in targeted studies. These findings could pave the way for the development of new biomarkers for early CVD detection and personalized prevention strategies. Future research should incorporate diverse cohorts and longitudinal sampling to deepen our understanding of the molecular mechanisms underlying CVD, enhance early disease prediction, and inform tailored interventions that improve patient outcomes.

## Data Availability

The Generation Scotland (GS) dataset is not publicly available as it contains information that could compromise participant consent and confidentiality. However, the data, research materials, and analytical methods will be made accessible to other researchers for the purpose of replicating the findings. Access will be granted upon successful project application to the GS Access Committee and obtaining ethical approval for accessing linked health data from NHS Scotland. Instructions for accessing GS data can be found at https://www.ed.ac.uk/generation-scotland/for-researchers/access; the GS Access Request Form can be downloaded from this site.

## Acknowledgments

The authors are grateful to all the families who took part, the general practitioners and the Scottish School of Primary Care for their help in recruiting them, and the whole Generation Scotland team, which includes interviewers, computer and laboratory technicians, clerical workers, research scientists, volunteers, managers, receptionists, health care assistants, and nurses.

## 7. Author contributions

R.E.M., and A.D.C. were responsible for the conception and design of the study. R.E.M. and A.D.C. drafted the article. A.D.C and D.L.M. carried out the data analyses. J.M., J.R, H.M.S contributed to the analyses and methodology. C.D. and H.G. were involved in investigation; sample preparation, collating approximately 10000 samples and transferring into required format for mass spectrometry analysis, QC checks and helping coordinate shipping. P.A., M.W., C.B.M, A.G., S.A., C.B., A.Z. and M.R. were responsible for mass spectrometry analysis. A.C. facilitated data linkage. C.H., M.R., A.Z, K.L.E. and J.F.P. were involved in conceptualisation and provided consultation on the methodology. D.J.P contributed to data collection and preparation. All authors read and approved the final manuscript.

## 8. Supplemental Material

Supplemental Methods Supplemental Tables 1-11

Supplemental Figures 1-8

Supplemental References 1-2

## 1. Non-standard Abbreviations and Acronyms

ABC: Ammonium Bicarbonate
ACN: Acetonitrile
BMI: Body Mass Index
BNP: B-type natriuretic peptide
CRP: C-reactive Protein
CVD: Cardiovascular Disease
DTT: Dithiothreitol
FA: Formic Acid
FDR: False Discovery Rate
GS: Generation Scotland
HCU: Hormonal Contraception Use
HR: Hazard Ratio
IAA: Iodoacetamide
LC: Liquid Chromatography
MS: Mass Spectrometry
NT: N-terminal
PG: Protein Group
PH: Proportional Hazards
SBP: Systolic Blood Pressure
SD: Standard Deviation
SIMD: Scottish Index of Multiple Deprivation
VIF: Variance Inflation Factor

