## Supplemental Methods for "Untargeted Proteomic Profiling Identifies Candidate Biomarkers for Early Detection of Cardiovascular Disease and Mortality"

**Protein Annotation**

Proteomics data were available for 15,818 Generation Scotland individuals. A total of 439 mass spectrometry proteins were classified as either individual proteins (n=133) or protein groups (n=306), based on the presence of dot (.) delimiters in their identifiers.

Protein group IDs were expanded into their component protein IDs (by splitting identifiers at the dot) and combined with individual protein IDs. The resulting dataset (n=596 protein IDs) was submitted to the UniProt ID Mapping tool (https://www.uniprot.org/id-mapping) for annotation. Retrieved protein names were pre-processed by removing text within parentheses and square brackets. Entries labelled as "deleted" in the annotation dataset were excluded from further analysis. Protein groups were classified as:

- Gene-derived protein groups (G): All proteins within a group shared at least one gene name.
- Mixed groups (PG): Proteins within a group originated from multiple distinct genes.

Gene-derived protein groups (G) were enumerated based on their recurrence (G1, G2, …), with single-instance groups labelled as G. Mixed groups were assigned sequential labels (PG 1, PG 2, …) and named based on the genes they include. For groups with four or fewer genes, all gene names are listed. For groups with more than four genes, a subset of gene names representing the diversity of the group is shown, followed by an ellipsis.

**Methodologies for Assessing Variables in Generation Scotland**

Age and sex variables were derived from clinical data cross-checked with genetic data (the presence of X and Y chromosomes). Systolic blood pressure was recorded as the average of two measurements. Laboratory analysis of blood samples collected from participants was conducted to measure levels of total cholesterol and HDL cholesterol. As triglyceride levels were not assessed in the Generation Scotland cohort, LDL cholesterol could not be derived; therefore, non-HDL cholesterol (total – HDL cholesterol) was used as a proxy. Disease status, educational attainment, contraceptive use, smoking status, and the number of cigarettes smoked per day were self-reported in a pre-clinic questionnaire.

Disease status was assessed by the question: "Please mark an X in the box if you, your father, mother, or any brother, sister, or grandparent has been affected by any of these conditions." Educational attainment was reported as the number of years spent attending school or studying full-time. As part of data post-processing, this variable was divided into 11 categories: 0) 0 years, 1) 1–4 years, 2) 5–9 years, 3) 10–11 years, 4) 12–13 years, 5) 14–15 years, 6) 16–17 years, 7) 18–19 years, 8) 20–21 years, 9) 22–23 years, 10) 24 or more years.

Females provided information about their use of contraceptives (contraceptive pill, injections, or implants), the age they started using contraception, and the duration of use. Contraceptive use was reported as a Boolean variable (“Have you ever taken the contraceptive pill or had contraceptive injections or implants?”). Age at which contraception started was categorized as follows: 1) <20, 2) 20–24, 3) 25–29, 4) 30–34, 5) 35–39, 6) 40–44, 7) 45–49, 8) 50–54, 9) 55–59, 10) 60+. Years of contraceptive use were divided into eight categories: 1) <1, 2) 1–2, 3) 3–4, 4) 5–9, 5) 10–14, 6) 15–19, 7) 20–24, 8) 25+.

Medication use was ascertained through linkage to Scottish National Health Service prescribing records. Prescriptions with British National Formulary codes beginning with 02 (cardiovascular system) were extracted and restricted to those issued within the six months prior to baseline.

Smoking status was classified as non-smoker, ex-smoker who stopped more than 12 months ago, ex-smoker who stopped within the past 12 months, and current smoker. Current and former smokers provided additional information, including the number of cigarettes smoked per day, age of smoking initiation, and age of smoking cessation (for former smokers only).

Lastly, data about participants’ residential addresses were used to assign a Scottish Index of Multiple Deprivation (SIMD) score.

**Derivation of Pack Years and Contraception Status**

The pack years variable was calculated by multiplying the duration of smoking (the difference between age of cessation and the age of initiation, in years) by the number of cigarettes smoked per day and then dividing the resulting number by 20 (the number of cigarettes in a pack). A value of zero was assigned to individuals who had never smoked.

Females were classified as currently using contraceptives if their current age fell within the time window that began at the age they started using contraception and ended at that age plus the duration of contraceptive use.

**Identifying Proteins Associated with Contraception Use**

As recent studies suggested a link between mass spectrometry data and contraception use ^1^, we explored this relationship in GS using ordinary least squares linear regression. The analysis considered only females (n=8,813). For each protein, its abundance was used as the response variable, with the use of contraceptive pills and age included as covariates. Models were fitted using the statsmodels ^2^ library in Python. To account for multiple testing, P values were adjusted using Bonferroni correction (P=0.05/439). Significant associations are reported in **Supplemental Table 2**.
