## Supplemental Figures for "Untargeted Proteomic Profiling Identifies Candidate Biomarkers for Early Detection of Cardiovascular Disease and Mortality"

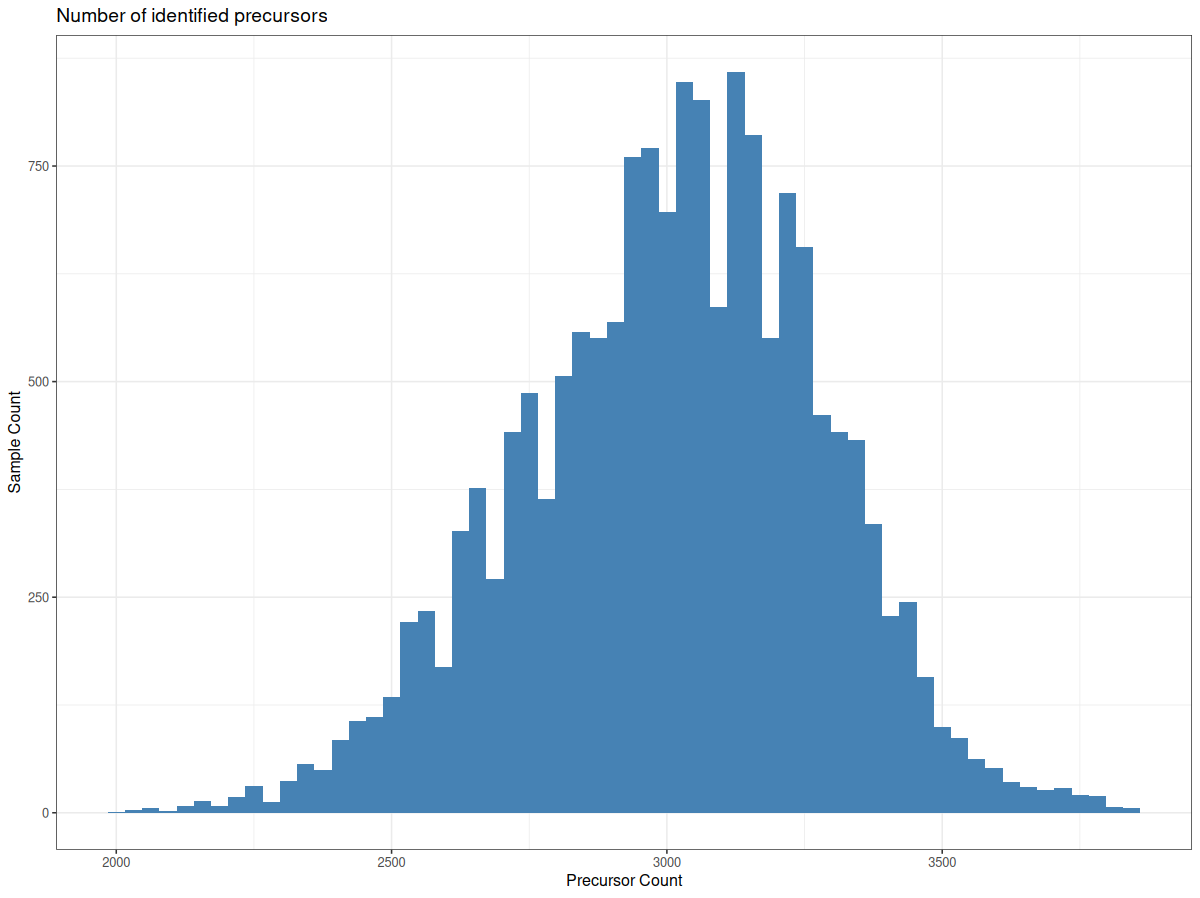

**Supplemental Figure 1.** Histogram of protein precursor counts**.**
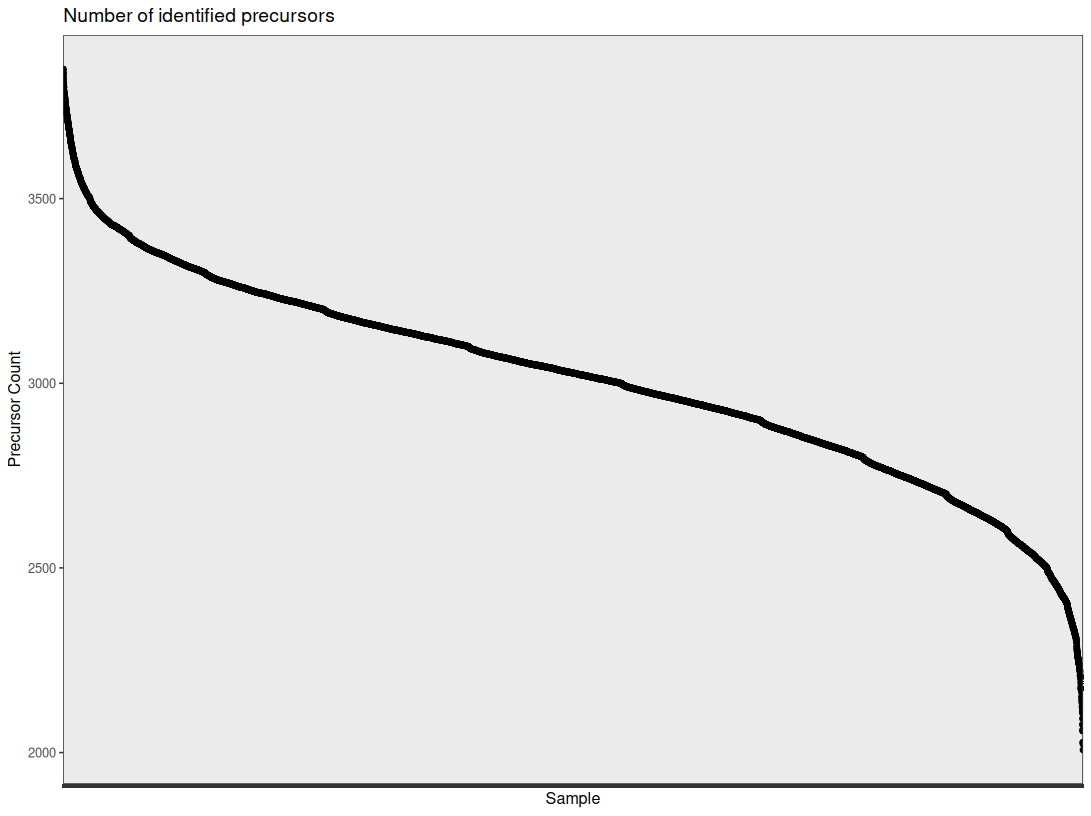

**Supplemental Figure 2.** Distribution of identified protein precursors across samples.

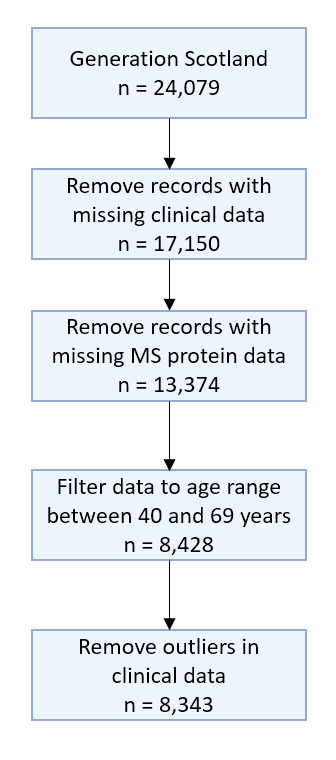

**Supplemental Figure 3.** Data cleaning pipeline. Clinical data included covariates of full models, such as age, sex, average systolic blood pressure, total cholesterol, HDL cholesterol, smoking status, presence of rheumatoid arthritis, diabetes status, years of education, and socioeconomic deprivation (measured by the Scottish Index of Multiple Deprivation). Average systolic blood pressure, log transformed pack years of smoking, HDL cholesterol and total cholesterol levels were trimmed of outliers (points beyond 4 standard deviations of the mean). BMI was log transformed and filtered to values between 18 and 50 kg/m^2^.

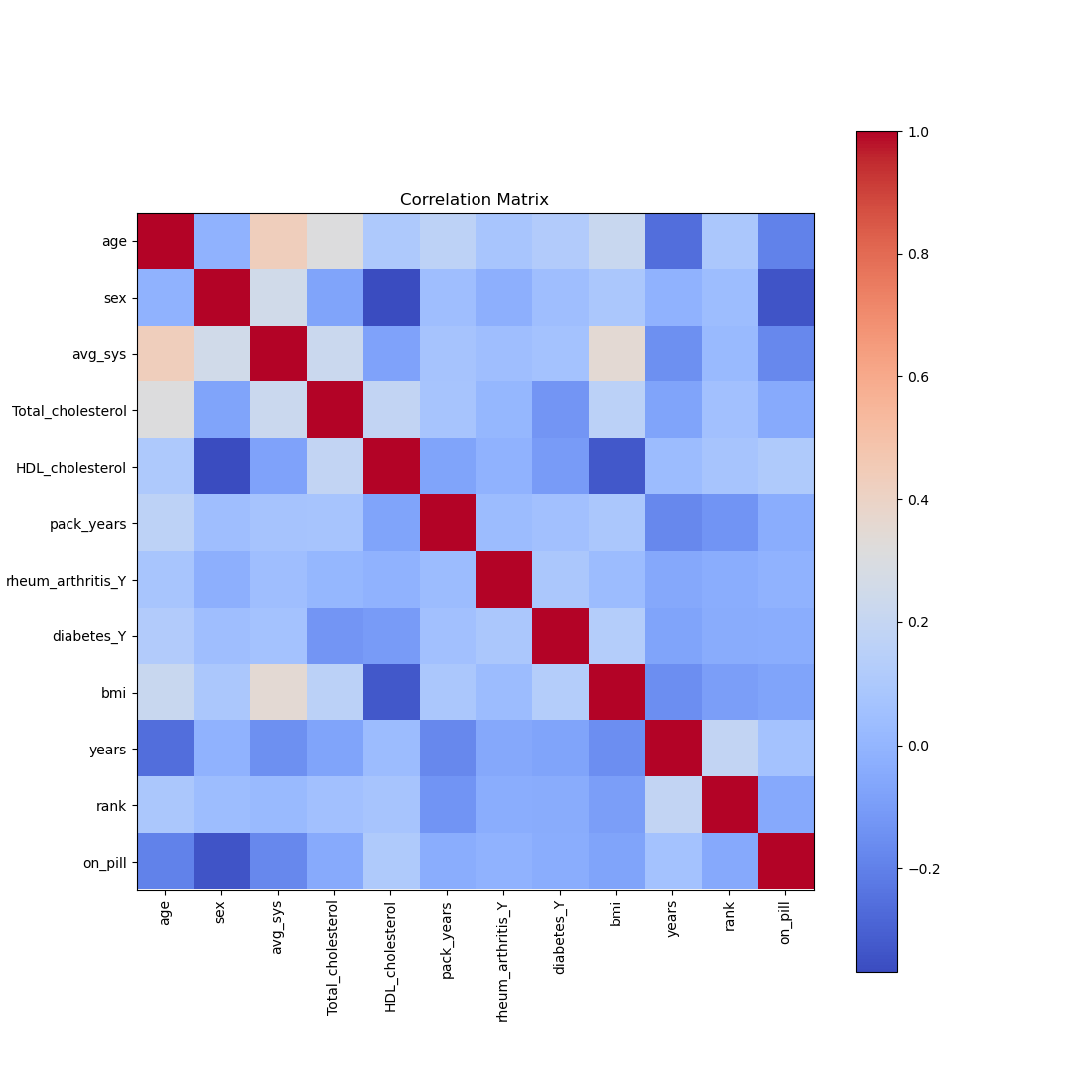

**Supplemental Figure 4.** Spearman correlation matrix displaying the correlations between coefficients of fully-adjusted Cox models. The abbreviations on the X and Y axes correspond to the following variables: avg_sys – average systolic blood pressure, rheum_arthritis_Y – rheumatoid arthritis status (yes/no), diabetes_Y – diabetes status (yes/no), bmi – Body Mass Index, years – years spent in full-time education, rank – SIMD score, on_pill – contraception use status (yes/no).

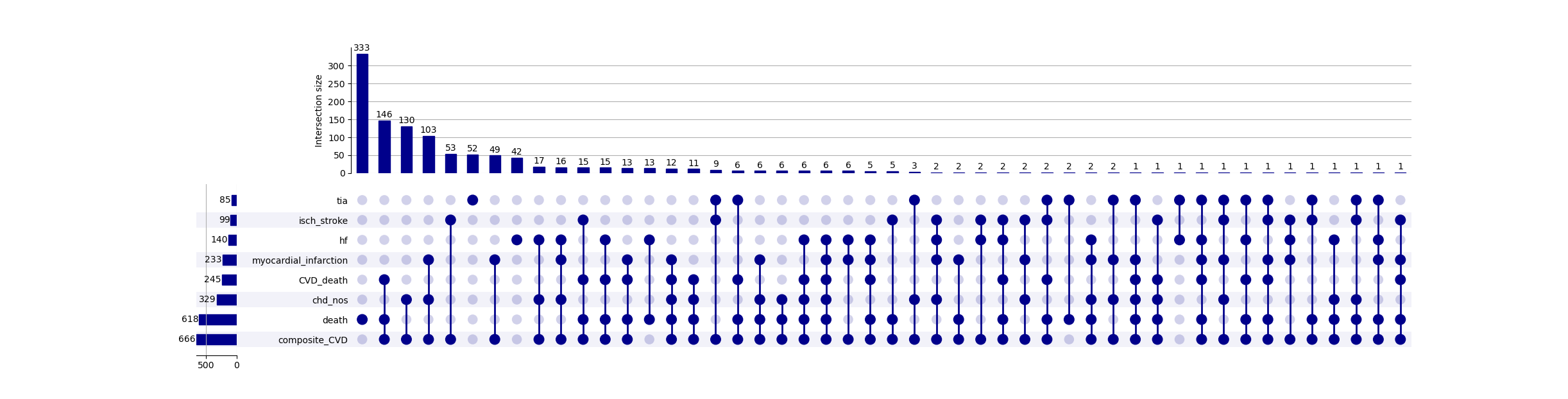

**Supplemental Figure 5**. Upset plot visualising intersections between event groups in Generation Scotland.

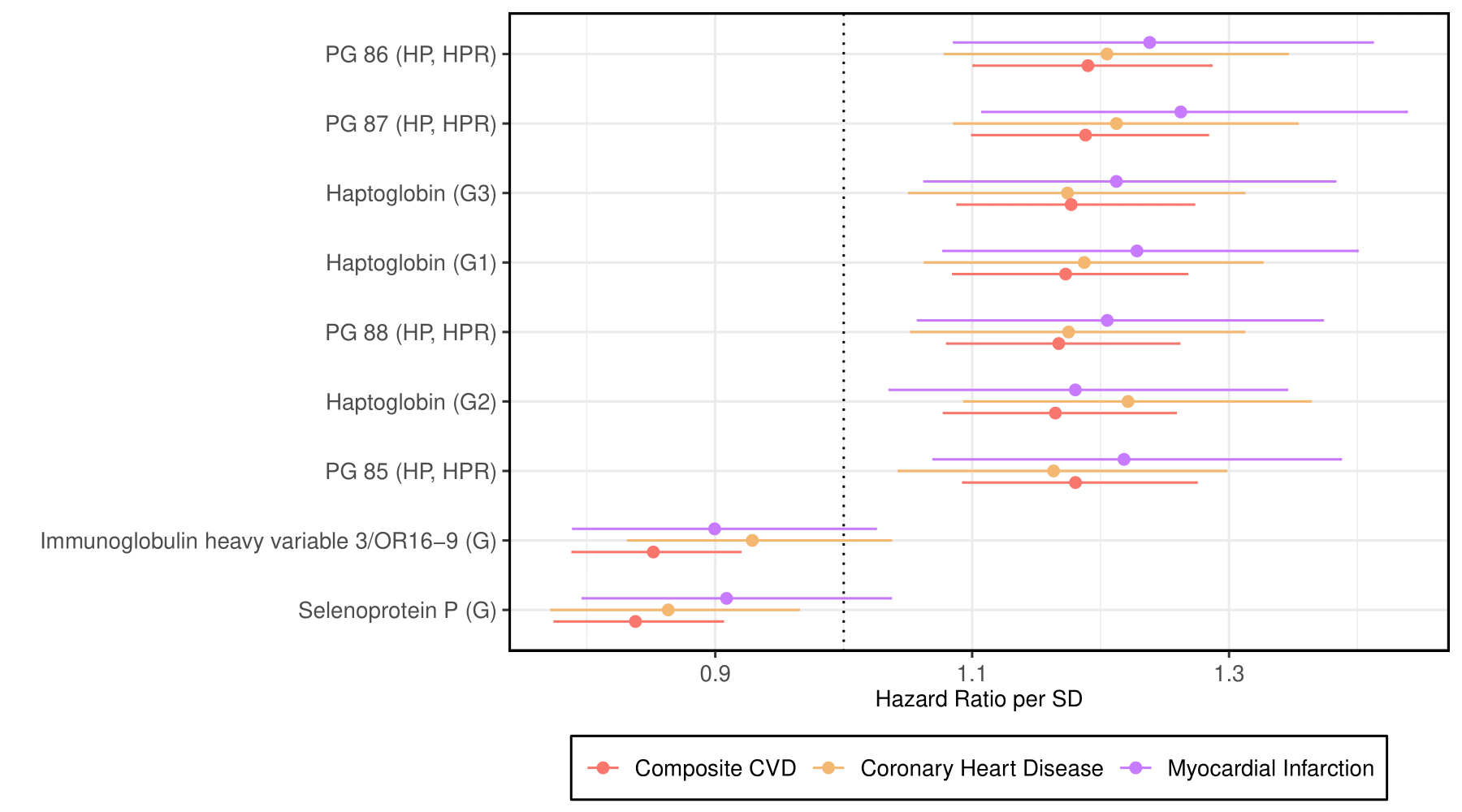

**Supplemental Figure 6.** Comparison of effect sizes and magnitudes between proteins significantly associated with either composite CVD or coronary heart disease or myocardial infarction in fully adjusted models (P < 0.05/439).

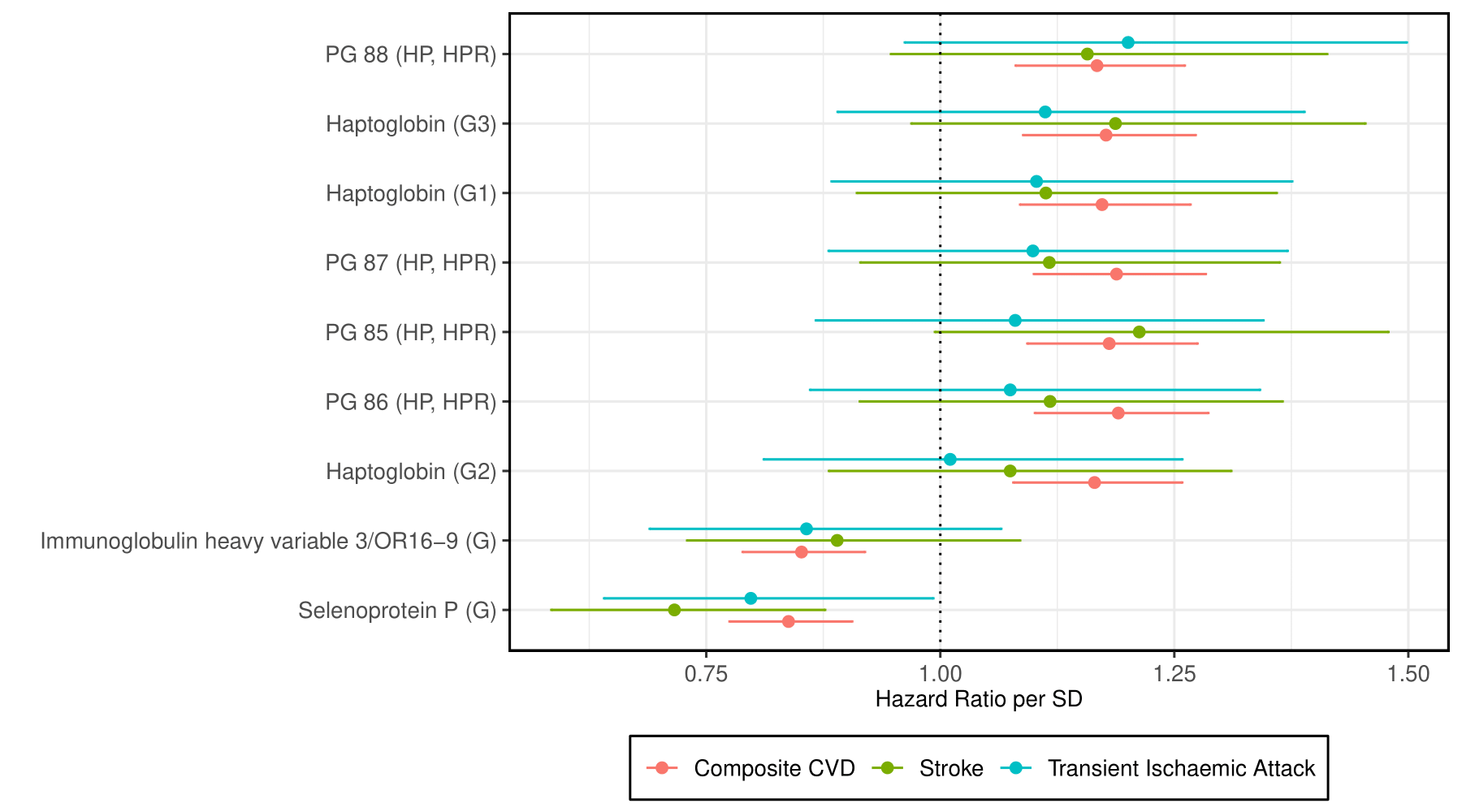

**Supplemental Figure 7.** Comparison of effect sizes and magnitudes between proteins significantly associated with either composite CVD or stroke or transient ischaemic attack in fully adjusted models (P < 0.05/439).

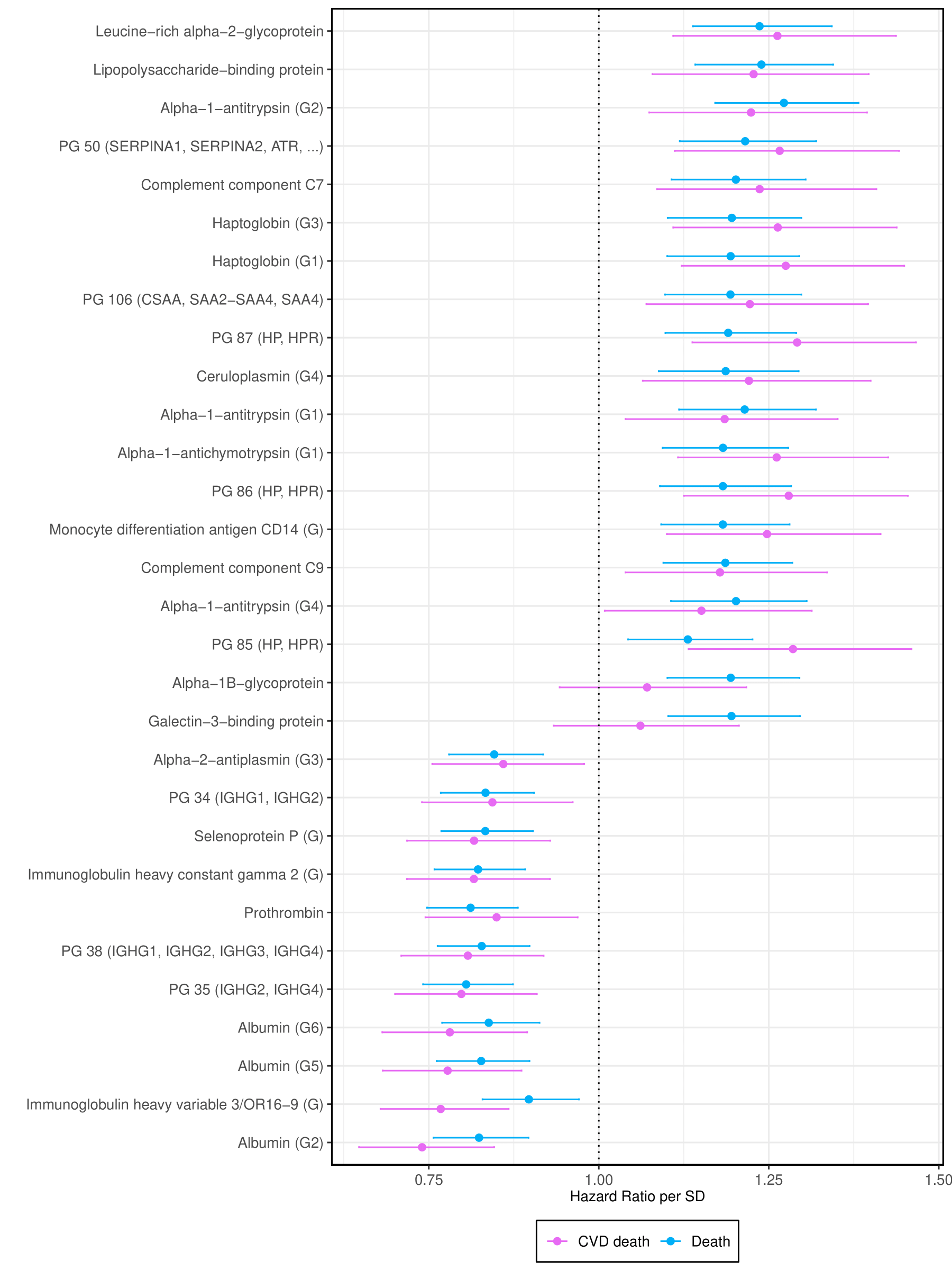

**Supplemental Figure 8.** Comparison of effect sizes and magnitudes between proteins significantly associated with either CVD death or all-cause death in fully adjusted models (P < 0.05/439).
